# The effect of serological screening for SARS-CoV-2 antibodies to participants’ attitudes and risk behaviour: a study on a tested population sample of industry workers in Split-Dalmatia County, Croatia

**DOI:** 10.1101/2020.06.15.20131482

**Authors:** Toni Ljubić, Ana Banovac, Ivan Buljan, Ivan Jerković, Željana Bašić, Ivana Kružić, Andrea Kolić, Rino Rivi Kolombatović, Ana Marušić, Šimun Anđelinović

## Abstract

Rapid serological tests for SARS-CoV-2 antibodies have been questioned by scientists and the public because of unexplored effects of negative test results on behaviour and attitudes, that could lower the level of adherence to protective measures. Therefore, our study aimed to investigate the changes in personal attitudes and behaviour before and after negative serological test results for SARS-CoV-2 antibodies. We conducted a survey questionnaire on 200 industry workers (69% males and 31% females) that have been previously tested negative. The survey examined participants’ self-reported general attitudes towards COVID-19, sense of fear, as well as their behaviour related to protective measures before and after the testing. The participants perceived the disease as a severe health threat and acknowledged the protective measures as appropriate. They reported a high level of adherence to measures and low level of fear both before and after the testing. Although those indicators were statistically significantly reduced after the test (P < 0.004), they did not result in risk behaviour. Therefore, the serological tests are not an additional threat regarding the risk behaviour in an environment where protective measures are efficient. In contrast, they might contribute to reducing the fear in the society and working environment.

## INTRODUCTION

Since November 2019, the Coronavirus (SARS-CoV-2) has been spreading around the globe leading to a pandemic with more than 4,5 million recorded cases and more than 300 000 deaths worldwide recorded on 15 May 2020 (Worldometer, 2020). Countries worldwide are testing their populations to estimate the number of people with active virus infection and the number of those who have recovered from it. It is highly recommended to prioritise testing hospitalised patients, healthcare facility workers, workers in congregate living settings, first responders, residents in long-term care facilities and generally persons with symptoms of (potential) COVID-19 infection using RT-PCR tests (Centers for Disease Control and Prevention, 2020). To estimate the number of people previously exposed to and/or infected with the virus, serological immunoassay tests are currently the best option, especially due to lower cost and shorter amount of time needed to obtain the results (Johns Hopkins Center For Health Security, 2020).

The first case of COVID-19 in the Republic of Croatia was reported in late February. To prevent the spreading and exponential growth of the disease, restrictive measures were introduced by the Croatian Government on 19 March 2020 (Koronavirus.hr, 2020a, 2020b). From 23 March 2020, leaving the place of residence was also prohibited (Koronavirus.hr, 2020c). With such restrictive measures, Croatia earned first place on the stringency scale provided by the Oxford COVID-19 Government Response Tracker on 26 March (Hale & Webster, 2020). Among mandatory protective measures, citizens were continuously provided with recommendations for personal protection, including social distancing (1 meter in open areas, 2 meters in closed areas), wearing face masks, maintaining personal hygiene, etc. Along with the national measures (Koronavirus.hr, 2020a, 2020b, 2020c), many companies introduced additional measures to protect the health of employees further and to maintain the manufacture. Such is the case for DIV Group, specialised in shipbuilding and production and trade of screws and mechanical parts, which introduced serological testing for employees using rapid serological immunoassay, as a health protection element within their corporate security system (DIV Group, 2020; Jerković et al., 2020).

Although the findings of serological testing for COVID-19 can be an essential part of investigating the disease, the tests vary in sensitivity and specificity and also produce false negative and false positive results (Long et al., 2020; West et al., 2020). These issues impose a danger to not only the health of tested individuals and communities but can also reduce the positive effects of national health policies and protective or restrictive measures necessary for the containment of the disease (Lippi et al., 2020). This can be especially devastating as some health experts worry that testing populations and providing them with a knowledge of their health and/or immunity status regarding COVID-19 could lead to yet unexplored psychological and behavioural effects (Green et al., 2020).

The aforementioned psychological and behavioural effects have already been investigated concerning negative test results received in various screenings. The concerns regarding these effects showed that people would, after receiving negative test results in a screening program, perceive they have a lower risk of developing the disease they were tested for and thus less likely take precautions not to get sick in future (Larsen et al., 2007; Marteau et al., 1996). A systematic review included eight studies on screening programs for diseases linked to lifestyle behaviours (type 2 diabetes, breast, bowel, lung and cervical cancer, and abdominal aortic aneurysm) to determine the post-screening changes in behaviours, attitudes, and emotions. The study showed that negative screening results are unlikely to cause changes in observed characteristics and to have a negative impact on behaviour (Cooper et al., 2017). Nevertheless, since COVID-19 is a novel disease whose spread is most effectively prevented by maintaining social distancing, community consciousness, personal protection and hygiene practices (Lakshmi Priyadarsini & Suresh, 2020) – behaviours that are most dependent on the conscientiousness and self-control of all individuals, it is of utmost importance to examine the behaviours and attitudes of people who received negative test results. Furthermore, these factors are vital in a specific working environment where interpersonal contact cannot be avoided entirely due to production characteristics. In these settings, changes in behaviour and attitudes of workers could impact the general psychological environment and, most importantly, the health of company workers and their families.

Thus, this study aims at investigating the changes in personal attitudes and behaviour of DIV Group industry workers before and after receiving negative serological test results for SARS-CoV-2 antibodies.

## METHODS

### Participants and setting

From May 10 to May 15, 2020, we conducted a survey of DIV Group industry workers in Split-Dalmatia County, Croatia, who were previously tested for SARS-CoV-2 antibodies by rapid immunoassays. The previous serological screening was conducted from April 23 to April 28, 2020, in collaboration with the Clinical Department for Pathology, Forensic Medicine and Cytology, University Hospital Centre Split and University Department of Forensic Sciences, University of Split (Split, Croatia). The named testing comprised 1316 participants and it was the first mass testing in the Republic of Croatia, and, to the authors’ knowledge, one of the first and largest studies on the corporative level in the world at that time (Jerković et al., 2020). That study analysed the test results of 1316 participants, revealing that only 0.99% of participants (95% CI 0.53–1.68) were positive for SARS-CoV-2 antibodies (Jerković et al., 2020).

The DIV Group facility in Split employs about 2200 people, which makes them the second largest employer in the county. The Split facility employee structure includes those working in production, as well as management and administration (DIV Group, 2020; Jerković et al., 2020).

To examine if the test results affected participants’ attitudes and behaviour, we constructed a short questionnaire and surveyed the employees, with the permission of the management of the company. The companies’ occupation safety officers distributed the questionnaire to the different company departments, including the management, administrative, and production staff. They visited different departments separately and offered employees that participated in the screening voluntary participation in the study. As only a small proportion (0.99%) of employees were tested positive for antibodies, we included only those with negative test results.

### Questionnaire

The questionnaire had six parts: (1) information of the study and informed consent; (2) general demographic data and test results; (3) participants’ general attitudes towards COVID-19; (4) participants’ protective behaviour and fear from the disease prior to testing; (5) participants’ protective behaviour and fear from the disease after the testing; and (6) the factors related to compliance with personal protection measures.

The general and demographic questions included gender, age, test results (negative / IgG positive/ IgM positive / IgM + IgG positive), and participants level of education. Other personal data were not included to ensure the participants’ anonymity.

The third part of the questionnaire included the questions on participants’ perception of the disease and its severity, as well as their attitudes on the protective and restrictive measurements given on the national and the company level. This question section provided seven statements that participants should have rated on a five-level Likert scale for agreement (1 = strongly disagree; to 5 = strongly agree).

In the fourth and the fifth part of the questionnaire, the participants were asked about their anxiety and fear of the COVID-19, compliance with restrictive measures and application of protective equipment before and after the testing. This section was composed of two sub-sections. The first one included nine statements regarding the participants’ fear and perception of their environment, that participants were asked to rate on a five-level Likert scale for agreement (1 = strongly disagree; to 5 = strongly agree). In the second sub-section, participants were asked to rate their frequency of obeying the restrictive measurements and applying the personal protective equipment. It included four statements with responses on a five-level Likert scale for frequency (1 = never; to 5 = very often).

In the final section, the participants were provided with four statements about factors that influence their adherence to the restrictive and protective measures, including the serological test results and level of actual restrictive measures and recommendations. They were also asked to select the one four statements that best suited their views.

The survey was approved by the University Department of Forensic Sciences Ethics Committee on 22 April 2020 (2181-227-05-12-19-0003; 024-04/19-03/00007). It was constructed in the Croatian language. The English translation is available in the supplementary material (Supplementary Material: Questionnaire).

### Statistical Analysis

Categorical variables, including the gender, education level, and factors affecting adherence to the protective measures, were given as frequencies and percentages. For the remaining variables, we provided the mean values with 95% confidence intervals. Differences in categorical variables were examined using the chi-squared test, while the differences in participants’ responses before and after the testing were examined using a paired-samples t-test. Due to the increased number of multiple comparisons (n=14), we set statistical significance at P ≤ 0.004 (Bonferroni correction). All analyses were performed with JASP 0.12.1 (JASP Team, 2020).

## RESULTS

The sample comprised 200 participants (68% men; median age = 43, interquartile range = 21). Most of them had undergraduate or graduate education (47.7%) or completed secondary education (32.7%), while fewer participants completed non-university college or professional studies (18.6%). There were only two participants with primary education (1%), and one answer was missing.

Most participants perceived COVID-19 as a dangerous disease and reported that restrictive measures and protective guidelines conducted on a national and company level were efficient and appropriate (Table 1).

**Table 1.**
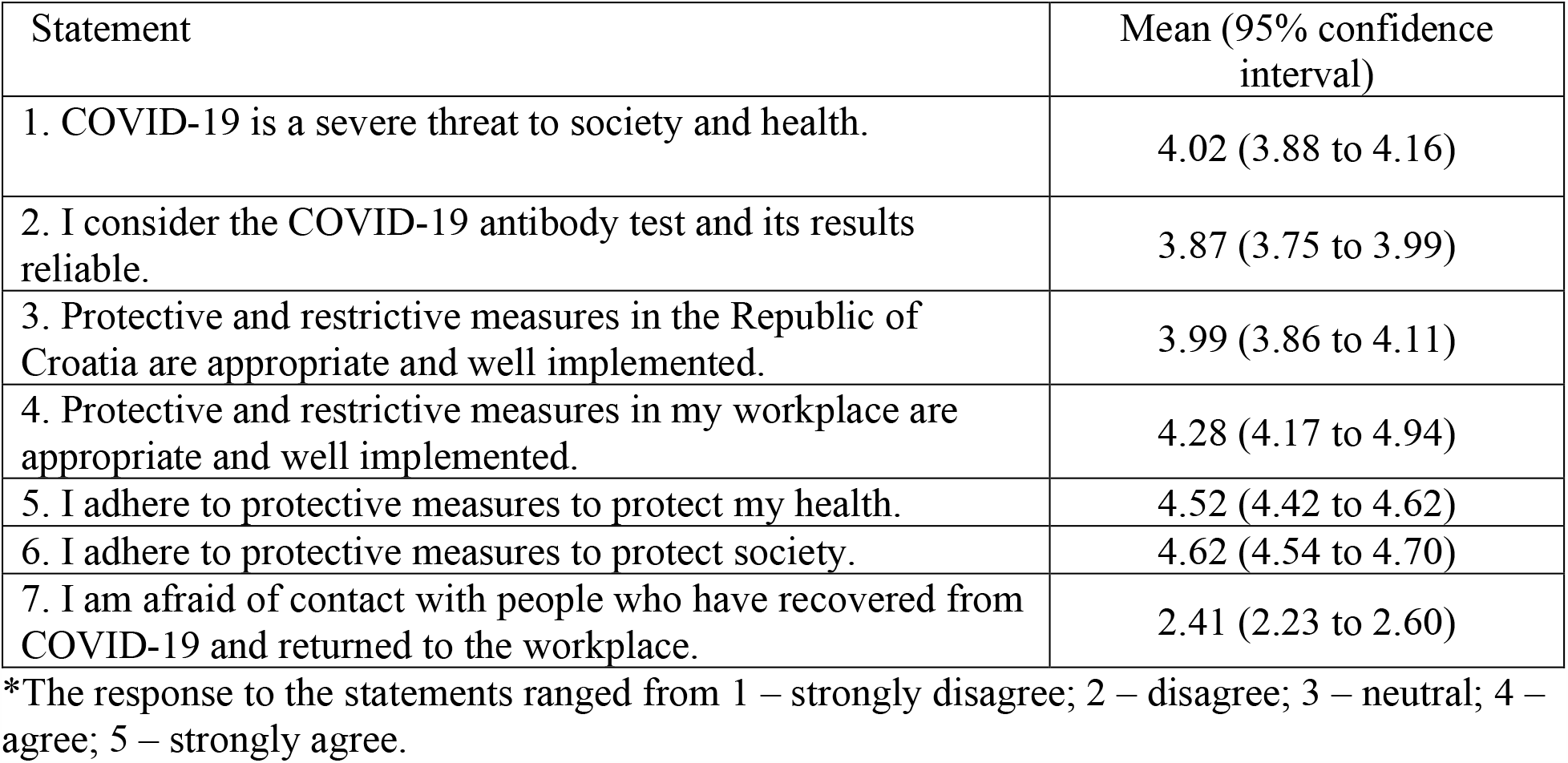
General attitudes on COVID-19 and protective measures*

| Statement | Mean (95% confidence interval) |
| --- | --- |
| 1. COVID-19 is a severe threat to society and health. | 4.02 (3.88 to 4.16) |
| 2. I consider the COVID-19 antibody test and its results reliable. | 3.87 (3.75 to 3.99) |
| 3. Protective and restrictive measures in the Republic of Croatia are appropriate and well implemented. | 3.99 (3.86 to 4.11) |
| 4. Protective and restrictive measures in my workplace are appropriate and well implemented. | 4.28 (4.17 to 4.94) |
| 5. I adhere to protective measures to protect my health. | 4.52 (4.42 to 4.62) |
| 6. I adhere to protective measures to protect society. | 4.62 (4.54 to 4.70) |
| 7. I am afraid of contact with people who have recovered from COVID-19 and returned to the workplace. | 2.41 (2.23 to 2.60) |

\*The response to the statements ranged from 1 – strongly disagree; 2 – disagree; 3 – neutral; 4 – agree; 5 – strongly agree.

On average, low levels of fear related to infection or infecting others with COVID-19 were observed both before and after the testing (Table 2, statements 1–6). Adherence to protective measures was also high prior to and post-testing (Table 2, statements 7–9). Nonetheless, changes in participants’ behaviour and attitudes before and after the testing were statistically significant for most variables. Suspicions and fear that a person or people in their physical vicinity were infected were significantly reduced. However, participants’ perception of other peoples’ adherence to measures did not change significantly (Table 2, statements 7–8).

**Table 2.** Participants’ self-reported behavioural characteristics before and after the test

| Statement | Mean (95% confidence interval) | Mean (95% confidence interval) | p* |
| --- | --- | --- | --- |
|  | Before the test | After the test |  |
| 1. I suspect I am infected with COVID-19. | 1.56 (1.43 to 1.70) | 1.33 (1.21 to 1.44) | **0.002** |
| 2. I suspect that some people in my work environment are infected with COVID-19. | 1.96 (1.81 to 2.14) | 1.61 (1.47 to 1.74) | **<0.001** |
| 3. I was afraid to stay with colleagues because I might infect them. | 2.10 (1.89 to 2.23) | 1.54 (1.41 to 1.68) | **<0.001** |
| 4. I was afraid to stay with colleagues because they might infect me. | 2.22 (2.05 to 2.38) | 1.72 (1.58 to 1.85) | **<0.001** |
| 5. I was afraid to stay with people who were not part of my household or immediate work environment because I might infect them. | 2.39 (2.20 to 2.57) | 1.69 (1.55 to 1.83) | **<0.001** |
| 6. I was afraid to stay with people who were not part of my household or immediate work environment because they might infect me. | 2.59 (2.40 to 2.77) | 2.19 (2.02 to 2.36) | **<0.001** |
| 7. People in my work environment have complied with all current protection measures. | 4.18 (4.10 to 4.30) | 4.13 (4.00 to 4.26) | 0.470 |
| 8. Members of my household and / or people I socialise with, adhered to all current protection measures to protect me. | 4.21 (4.07 to 4.35) | 4.11 (3.97 to 4.25) | 0.145 |
| 9. Members of my household and / or people I socialise with, adhered to all current protection measures to protect others. | 4.29 (4.16 to 4.41) | 4.10 (3.96 to 4.24) | **0.002** |
| Total score | 25.43 (24.57 to 26.29) | 22.39 (21.66 to 23.12) | **<0.001** |

The response to the statements ranged from 1 – strongly disagree; 2 – disagree; 3 – neutral; 4 – agree; 5 – strongly agree.
\*Paired-samples t-test.
†Statistically significant values are in bold.

The participants on average showed a high frequency of adherence to protective measures and restrictions (Table 3). When they were asked about their pre-test and post-test adherence frequencies, they reported maintaining the application of personal protective equipment on the almost same level, but lower adherence to social distancing (Table 3, statements 2–4).

**Table 3.** The self-reported frequency of participants adherence to protective measures

| Statement | Mean (95% confidence interval) | Mean (95% confidence interval) | P* |
| --- | --- | --- | --- |
|  | Before the test | After the test |  |
| 1. Since the introduction of restrictive measures, I have worn protective equipment (mask and / or gloves). | 4.29 (4.17 to 4.40) | 4.17 (4.04 to 4.29) | 0.011 |
| 2. Since the introduction of restrictive measures, I have maintained a social distance. | 4.46 (4.36 to 4.56) | 4.29 (4.18 to 4.40) | **<0.001** |
| 3. Since the introduction of restrictive measures, I have avoided socialising with more than 5 people. | 4.51 (4.40 to 4.62) | 4.19 (4.05 to 4.32) | **<0.001** |
| 4. Since the introduction of restrictive measures, I have avoided socialising with people who are not part of my household and immediate work environment. | 4.32 (4.19 to 4.44) | 4.00 (3.89 to 4.14) | **<0.001** |
| Total score | 17.57 (17.20 to 17.94) | 16.64 (16.21 to 17.07) | **<0.001** |

The response to the statements ranged from 1 – never; 2 – seldom; 3 – sometimes; 4 – frequently; 5 – very frequently.
\*Paired-samples t-test.
†Statistically significant values are in bold.

Although the participants reported changes in behaviour and attitudes before and after receiving the test results, most of the participants did not attribute their behaviour to the test itself but rather to the level of company and national protective measures (Table 4).

**Table 4.**
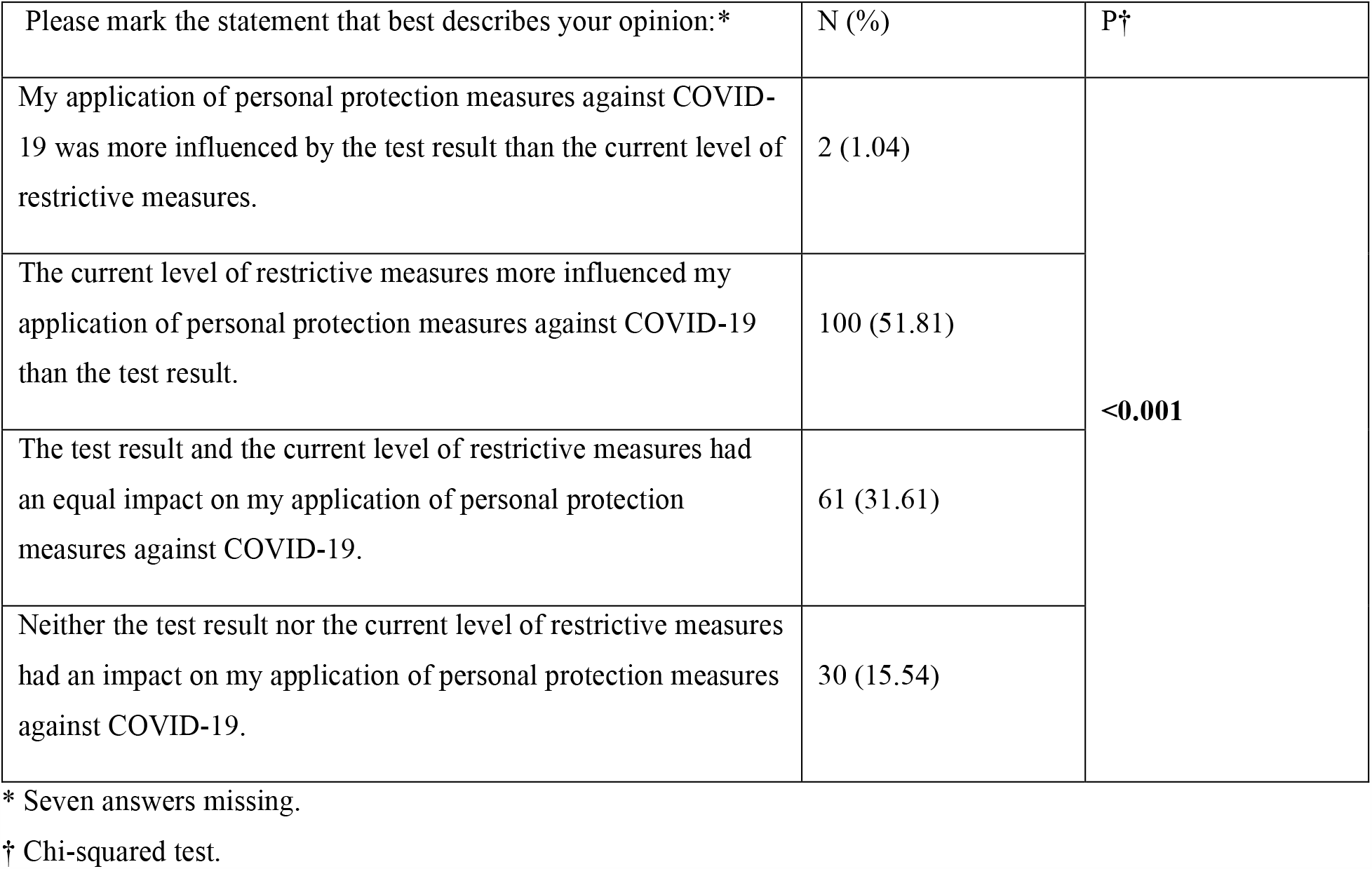
Factors that affected the participants’ adherence to protective measures.

| Please mark the statement that best describes your opinion:* | N (%) | P† |
| --- | --- | --- |
| My application of personal protection measures against COVID-19 was more influenced by the test result than the current level of restrictive measures. | 2 (1.04) | **<0.001** |
| The current level of restrictive measures more influenced my application of personal protection measures against COVID-19 than the test result. | 100 (51.81) |  |
| The test result and the current level of restrictive measures had an equal impact on my application of personal protection measures against COVID-19. | 61 (31.61) |  |
| Neither the test result nor the current level of restrictive measures had an impact on my application of personal protection measures against COVID-19. | 30 (15.54) |  |

\* Seven answers missing.
† Chi-squared test.

## DISCUSSION

The results of the present study showed that negative serological test result is associated with changes in the behaviour and attitudes of participants, but not to the extent that would lead to irresponsible or dangerous behaviour. To the best of our knowledge, this is the first study that investigates the changes before and after receiving negative serological COVID-19 test results on behaviours and attitudes connected to this disease.

The results of this study indicate that the levels of fear of being infected or infecting others with COVID-19, as well as behaviours regarding adherence to protective measure, changed significantly in the timeframe after receiving negative test results. However, the subjects’ fear of infection and/or infecting was initially at the lower moderate to low level and dropped to an even lower level after the testing. Although the disease had a pandemic character and was at that time relatively unexplored, the situation at the company was under control as the company introduced protective measures at the end of February. This was influenced by the experience of their partners in China and Italy, which were at the time global pandemic hotspots. These measures, along with national protective measures, introduced by the second half of March (Koronavirus.hr, 2020a, 2020b, 2020c), were probably beneficial for the participants’ lower level of fear.

The frequency of positive behaviour related to social distancing also reduced after the testing, but still remained high. In contrast, results indicate no significant change in behaviours related to wearing protective equipment, masks, and gloves which were highly adherent. Both of the findings could be attributed to the stimulating climate in the company and society that raised awareness of protective and restrictive measures. We also did not find changes in the perception of colleagues’ compliance with protective measures pre- and post-testing. These findings might additionally support the participants’ responsibility and conscientiousness, regardless of their test results. This is also evident from the fact that most of them attributed their behaviour less to the tests results but more to the current level of restrictive measures.

Studies on screening for various diseases such as different types of cancer, sexually transmitted diseases (STDs), diabetes, etc. have been conducted to determine their psychological and behavioural effects but also the perception of ones’ health and future risk of getting sick (Ashraf et al., 2009; Berstad et al., 2015; Collins et al., 2011; Eborall et al., 2007; Sznitman et al., 2010). The recent review on these types of studies shows a small decrease in perceived risk of the disease screened for, slightly lower levels of anxiety or worry in the screen negative group, and highlights that out of 28 studies only five showed an unfavourable change in the negatively screened groups’ health-related behaviours (Cooper et al., 2017).

Although our study findings indicate changes of similar direction and extent, it is difficult to compare its results to abovementioned studies. This is due to the very nature of COVID-19, an infectious disease spread primarily by human contact and interaction. The other diseases populations are usually screened for are (Ashraf et al., 2009; Berstad et al., 2015; Collins et al., 2011; Eborall et al., 2007; Sznitman et al., 2010), except for STDs, not transmittable. But the comparison is not possible even with STDs since their transmittance is usually restricted to the most intimate of human interactions and thus limited. The cessation of infectious disease spreading such as COVID-19 is impossible without necessary changes in human interactions and behaviour, which must be applied to all members of society.

The limitation of this study is that compared pre- and post-testing self-ratings were all given after the testing, thus potentially introducing a reporting bias. To obtain pre-testing measurements, it was only possible to survey the participants on the day of voluntary serological testing. From the organisational and protective standpoint, it was of utmost importance to minimise the time participants spent at the testing station to the time required for serological testing and completing a mandatory accompanying questionnaire on disease-related factors (Jerković et al., 2020). This would result in not only the prolonged absence of participants from their workplace but also their potentially increased exposure to the virus. Since the period between testing and completing the survey questionnaire lasted a maximum of 21 days for each participant, by testing at a single point in time we relied on the participants’ ability to recall recent behaviours and attitudes. While other studies on the impact of negative screening results repeated measurements after several months or years (Cooper et al., 2017), this was not possible for this study due to the very nature of COVID-19 as well as differing levels of national restrictive measures.

The additional limitation of this study was the lack of a control group. The COVID-19 screening in DIV Group in Split (Jerković et al., 2020) resulted in an insufficient number of positive participants to represent a separate group of subjects in research. Therefore, due to the extremely low seroprevalence in the tested sample (about 1%), including positive participants would not provide relevant information for the scope of the study. Also, having an adequate control group of non-tested participants was not possible since almost all DIV Group industry workers in Split were screened. Surveying the general population for that purpose would not be appropriate, as DIV employees were immersed in an all-encompassing working atmosphere with special and more severe protection measures prescribed by the employer, that were introduced considerably earlier than the national measures. However, even if we would have detected that general population control group had adhered less to the protective measures, due to social climate influenced by the smaller number of newly infected or the current level of national restrictions, it could have only implied that test results had even fewer negative consequences on behaviour related to protective measures.

In conclusion, our study results indicate that COVID-19 serological testing does not impose an additional threat regarding the potentially irresponsible or risk behaviour in an environment where protective measures are efficient. Additionally, they might be beneficial to reducing the level of fear in society and the working environment.

## Data Availability

The data that support the findings of this study are available from the corresponding author, IJ, upon reasonable request.

## DISCLAIMER

This article does not represent in whole or in part the views of the authors’ institutions. However, it does express those of the authors.

## ACKNOWLEDGEMENTS

The authors would like to thank DIV Group company and Tomislav Debeljak, along with all study participants. We are especially thankful to Boško Ramljak, Marija čečuk and Ivica Sinovčić for their assistance in organisation and data collection.

---

Dear participant,

In front of you is a 4-page questionnaire which aims to examine the impact of the results of serological testing for SARS-CoV antibodies on personal attitudes and behaviour of participants. This survey is conducted by the University Department of Forensic Sciences in cooperation with the Medical Faculty of the University of Split and the Clinical Department of Pathology, Forensic Medicine and Cytology of the Clinical Hospital Centre Split. The research was approved by the ethics committee of the University Department of Forensic Sciences (2181-227-05-12-19-0003; 024-04 / 19-03 / 00007). <u>The survey is conducted anonymously, and your personal data will not be available to researchers or your employer, and the results will be used exclusively for research purposes.</u>

It takes a maximum of 10 minutes to complete the questionnaire.

**Do you agree to participate in this research?**

**YES**

**NO**

**Circle your gender:** M / F

**Write down your age (in years):____**

**Circle your test result:** negative / IgG positive / IgM positive / IgM + IgG positive

**Circle the highest completed level of your education:**

primary education

non-university college or professional studies

secondary education

undergraduate or graduate education

For the following statements, please circle your **level of agreement** (one answer for each statement).

1. *strongly disagree*
2. *disagree*
3. *neither agree nor disagree*
4. *agree*
5. *strongly agree*

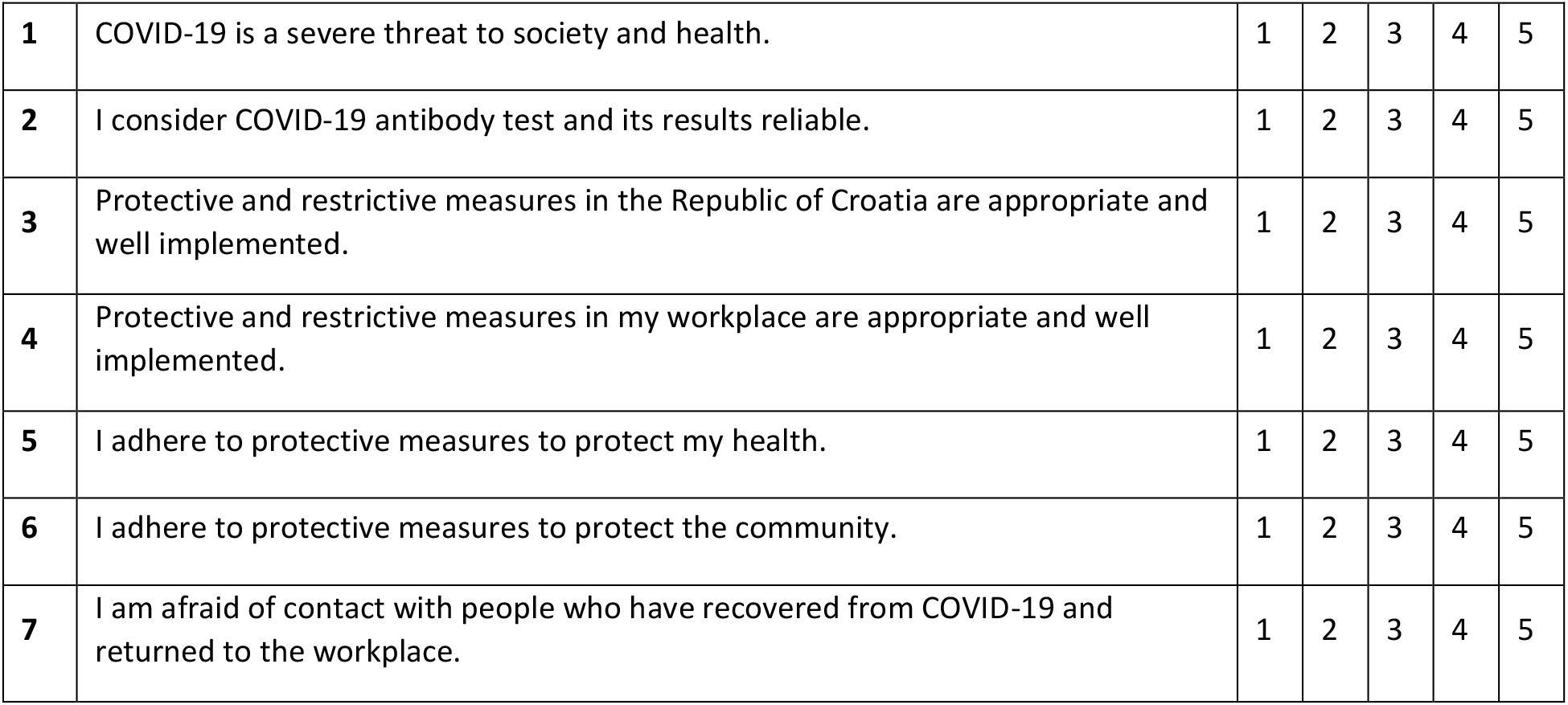

For the following statements, please circle your **level of agreement** (one answer for each statement).

1. *strongly disagree*
2. *disagree*
3. *neither agree nor disagree*
4. *agree*
5. *strongly agree*

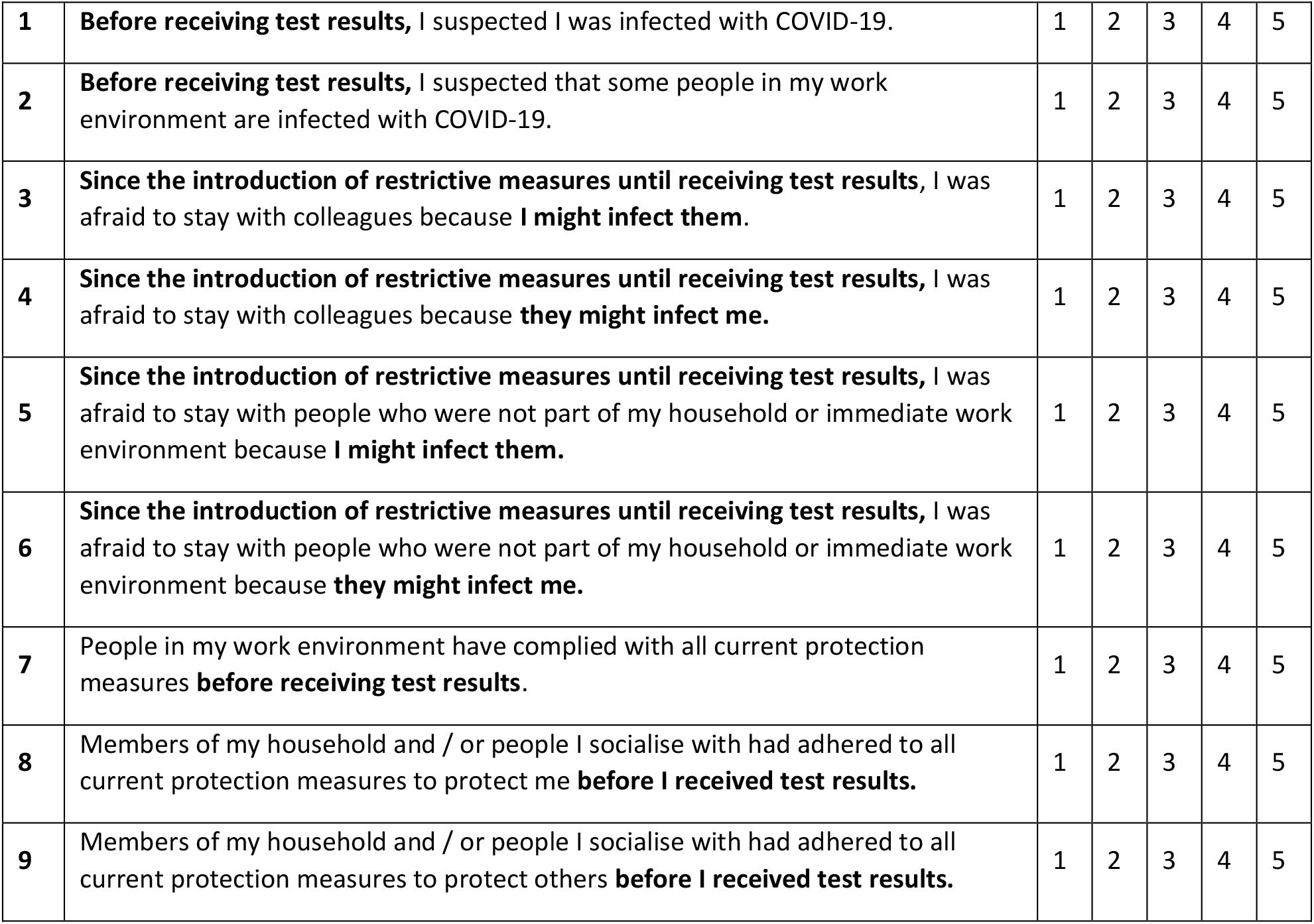

|  |  |  |  |  |  |  |
| --- | --- | --- | --- | --- | --- | --- |
| **1** | **Before receiving test results**, I suspected I was infected with COVID-19. | 1 | 2 | 3 | 4 | 5 |
| **2** | **Before receiving test results**, I suspected that some people in my work environment are infected with COVID-19. | 1 | 2 | 3 | 4 | 5 |
| **3** | **Since the introduction of restrictive measures until receiving test results**, I was afraid to stay with colleagues because **I might infect them**. | 1 | 2 | 3 | 4 | 5 |
| **4** | **Since the introduction of restrictive measures until receiving test results**, I was afraid to stay with colleagues because **they might infect me**. | 1 | 2 | 3 | 4 | 5 |
| **5** | **Since the introduction of restrictive measures until receiving test results**, I was afraid to stay with people who were not part of my household or immediate work environment because **I might infect them**. | 1 | 2 | 3 | 4 | 5 |
| **6** | **Since the introduction of restrictive measures until receiving test results**, I was afraid to stay with people who were not part of my household or immediate work environment because **they might infect me**. | 1 | 2 | 3 | 4 | 5 |
| **7** | People in my work environment have complied with all current protection measures **before receiving test results**. | 1 | 2 | 3 | 4 | 5 |
| **8** | Members of my household and / or people I socialise with had adhered to all current protection measures to protect me **before I received test results**. | 1 | 2 | 3 | 4 | 5 |
| **9** | Members of my household and / or people I socialise with had adhered to all current protection measures to protect others **before I received test results**. | 1 | 2 | 3 | 4 | 5 |

For the following statements, please circle your **level of frequency** (one answer for each statement).

1. *never*
2. *rarely*
3. *sometimes*
4. *often*
5. *always*

|  |  |  |  |  |  |  |
| --- | --- | --- | --- | --- | --- | --- |
| **1** | **Since the introduction of restrictive measures until receiving test results**, I have worn protective equipment (mask and / or gloves): | 1 | 2 | 3 | 4 | 5 |
| **2** | **Since the introduction of restrictive measures until receiving test results**, I have maintained a social distance: | 1 | 2 | 3 | 4 | 5 |
| **3** | **Since the introduction of restrictive measures until receiving test results**, I have avoided socialising with more than 5 people: | 1 | 2 | 3 | 4 | 5 |
| **4** | **Since the introduction of restrictive measures until receiving test results**, I have avoided socialising with people who are not part of my household and immediate work environment: | 1 | 2 | 3 | 4 | 5 |

For the following statements, please circle your **level of agreement** (one answer for each statement).

1. *strongly disagree*
2. *disagree*
3. *neither agree nor disagree*
4. *agree*
5. *strongly agree*

|  |  |  |  |  |  |  |
| --- | --- | --- | --- | --- | --- | --- |
| **1** | I suspected I was infected with COVID-19 **after receiving test results.** | 1 | 2 | 3 | 4 | 5 |
| **2** | **Before receiving test results**, I suspected that some people in my work environment are infected with COVID-19 after receiving test results. | 1 | 2 | 3 | 4 | 5 |
| **3** | **After receiving the test results**, I was afraid to stay with colleagues because **I might infect them.** | 1 | 2 | 3 | 4 | 5 |
| **4** | **After receiving the test results**, I was afraid to stay with colleagues because **they might infect me.** | 1 | 2 | 3 | 4 | 5 |
| **5** | **After receiving test results**, I was afraid to stay with people who were not part of my household or immediate work environment because **I might infect them.** | 1 | 2 | 3 | 4 | 5 |
| **6** | **After receiving test results**, I was afraid to stay with people who were not part of my household or immediate work environment because **they might infect me.** | 1 | 2 | 3 | 4 | 5 |
| **7** | People in my work environment have complied with all current protection measures **after receiving test results.** | 1 | 2 | 3 | 4 | 5 |
| **8** | Members of my household and / or people I socialise with have adhered to all current protection measures to protect me **after I received test results.** | 1 | 2 | 3 | 4 | 5 |
| **9** | Members of my household and / or people I socialise with have adhered to all current protection measures to protect others **after I received test results.** | 1 | 2 | 3 | 4 | 5 |

For the following statements, please circle your **level of frequency** (one answer for each statement).

1. *never*
2. *rarely*
3. *sometimes*
4. *often*
5. *always*

|  |  |  |  |  |  |  |
| --- | --- | --- | --- | --- | --- | --- |
| **1** | **After receiving test results**, I have worn protective equipment (mask and / or gloves): | 1 | 2 | 3 | 4 | 5 |
| **2** | **After receiving test results**, I have maintained a social distance: | 1 | 2 | 3 | 4 | 5 |
| **3** | **After receiving test results**, I have avoided socialising with more than 5 people: | 1 | 2 | 3 | 4 | 5 |
| **4** | **After receiving test results**, I have avoided socialising with people who are not part of my household and immediate work environment: | 1 | 2 | 3 | 4 | 5 |

**Circle the statement that is most accurate for you:**

1. My application of personal protection measures against COVID-19 was more influenced by the test result than the current level of restrictive measures.
2. My application of personal protection measures against COVID-19 was more influenced by the current level of restrictive measures than by the test result.
3. The test result and the current level of restrictive measures had an equal impact on my application of personal protection measures against COVID-19.
4. Neither the test result nor the current level of restrictive measures had an impact on my application of personal protection measures against COVID-19.

> Thank you for participating!

